# Temporal Variability of the Electromechanical Window in Long-QT Syndrome and Drug-Induced QT Prolongation: Value for Enhanced Arrhythmia-Risk Assessment

**DOI:** 10.1101/2025.09.03.25334690

**Authors:** Peter M. Deissler, Ann-Kathrin Rahm, Anat Berkovitch, Mara Elena Müller, Maurits Sikking, Stephane Heymans, Benedikt Langenberg, Maximilian Moersdorf, Marina Rieder, Saranda Nimani, Katja E. Odening, Wolfgang Dichtl, Avi Sabbag, Paul G. A. Volders, Rachel M.A. ter Bekke

**Author notes:** Address of correspondence: Dr. Rachel ter Bekke, MD PhD Department of Cardiology, Cardiovascular Research Institute Maastricht, Maastricht University Medical Center+, P.O. Box 5800, 6202 AZ, Maastricht, The Netherlands. There is no conflict of interest.

## Abstract

**Background:** Arrhythmia-risk assessment in congenital long-QT syndrome (LQTS) and drug-induced QT prolongation (diQTP) is primarily based on clinical, genetic, and electrical parameters. Electromechanical window (EMW; aortic-valve closure time minus QT interval) assessment outperformed QTc as a predictor of symptomatic status in LQTS. The relationship between QTc and EMW dynamics, and ventricular tachyarrhythmia (VT) timing in LQTS and diQTP is unknown.

**Methods:** 47 LQTS/-VT, 18 LQTS/+VT (3 LQT1, 5 LQT2, 4 LQT3, 3 LQT7, 3 genotype-negative), nine diQTP/+VT and 26 controls were included. QTc and EMW were obtained from standard 12-lead ECGs and ECG-echocardiograms at two or three time points. +VT patients were included if EMW/QTc assessments were performed within two weeks before or after torsades de pointes, ventricular fibrillation, monomorphic or bidirectional VT.

**Results:** Median age was 42 (27-57) years, and 70% were female. In control subjects, EMW remained stably positive over time. In LQTS/-VT patients, EMW was negative without significant variation. In LQTS/+VT and diQTP/+VT patients, transient accentuations of EMW negativity were observed at the time point closest to VT (2 (1 to 7) days to arrhythmia), regardless of whether measured before or after VT. Temporary EMW negativity accentuation was driven by foreshortening of mechanical systole despite concurrent QT prolongation. EMW recovery after VT was similar for patients with or without beta-blocker therapy. Multiple logistic regression analysis identified EMW negativity and EMW dynamics (ΔEMW) as independent predictors of imminent VT in LQTS. An EMW of −75 ms and ΔEMW of −39 ms were optimal cut-offs to predict emergent arrhythmic deterioration in the LQTS cohort.

**Conclusion:** Temporary accentuation of EMW negativity is a marker of impending VT in LQTS and diQTP patients. EMW negativity ≤ −75 ms and a ΔEMW ≥ −39 ms best differentiated LQTS patients with imminent or very recent VT from those not at acute risk.

## Introduction

Sudden cardiac death (SCD) accounts for up to 50% of all cardiac deaths and is primarily caused by ventricular tachyarrhythmias (VT) and fibrillation (VF).^1^ Inherited channelopathies, such as congenital long-QT syndrome (LQTS), account for 5–10% of SCD cases; however, the annual arrhythmia risk in LQTS patients is relatively low at 0.47%.^1–3^ Drug-induced QT prolongation (diQTP) occurs more commonly and can be found in ∼6% of patients treated with repolarization-delaying cardiac or non-cardiac compounds. Less than 5% of diQTP patients develop VT.^4^

Risk stratification in LQTS is primarily based on ECG characteristics (prolongation of heart rate-corrected QT interval (QTc) and T-wave dynamics), torsades de pointes (TdP), syncope, sex, genotype, and family history.^5–7^ Serial ECG assessment may further enhance risk stratification, as QTc values in LQTS patients fluctuate over time.^8,9^

The electromechanical window (EMW) is an ECG-echocardiographic parameter that indicates the relation between repolarization duration and left-ventricular contraction duration.^10^ Previous studies have demonstrated that a single measurement of EMW outperforms the heart rate-corrected QT interval (QTc) in predicting the symptomatic status of LQTS patients.^10–12^ LQTS patients with a remote history of sudden cardiac arrest or TdP had a more negative EMW than those without, even for the same QT interval.^10–12^ Also, in a case report, profound EMW negativity was associated with life-threatening arrhythmic events in diQTP.^13^ Whether accentuated EMW negativity in LQTS/diQTP patients reflects an increased imminent risk of VT remains unknown. Here, we investigate the temporal variability of EMW in a large group of LQTS and diQTP patients with documented VT, hypothesizing that EMW is more negative during episodes of electrical instability and arrhythmia. In addition, we examine whether accentuated EMW negativity and its change over time (ΔEMW) are associated with the timing of VT, to assess their potential for improving arrhythmia-risk prediction.

## Methods

### Study Design and Criteria

Subjects were enrolled retrospectively from six centers across five countries, with written informed consent obtained following the Declaration of Helsinki. Control subjects were recruited after cardiac abnormalities were excluded. LQTS was diagnosed according to the ESC Guidelines for the Management of Patients with Ventricular Arrhythmias and the Prevention of Sudden Cardiac Death,^1^ and diQTP as transient QT prolongation during treatment with known repolarization-delaying drugs (https://crediblemeds.org/). Arrhythmic events (VT) were defined as documented TdP, VT, or VF. LQTS/diQTP+VT patients were only included if echocardiography was performed less than 14 days before or after VT (Figure 1A). ECG and echocardiographic characteristics were assessed at two or three time points. According to this study design, the reference time point was set as the first time point for controls and LQTS/-VT, and the time point closest to VT for LQTS/+VT and diQTP/+VT. Parameter changes between time points (Δ) were calculated as mean of change between available time points (see results section for details). Data will be shared by the corresponding author upon reasonable request and after legal assessment.

**Figure 1:**
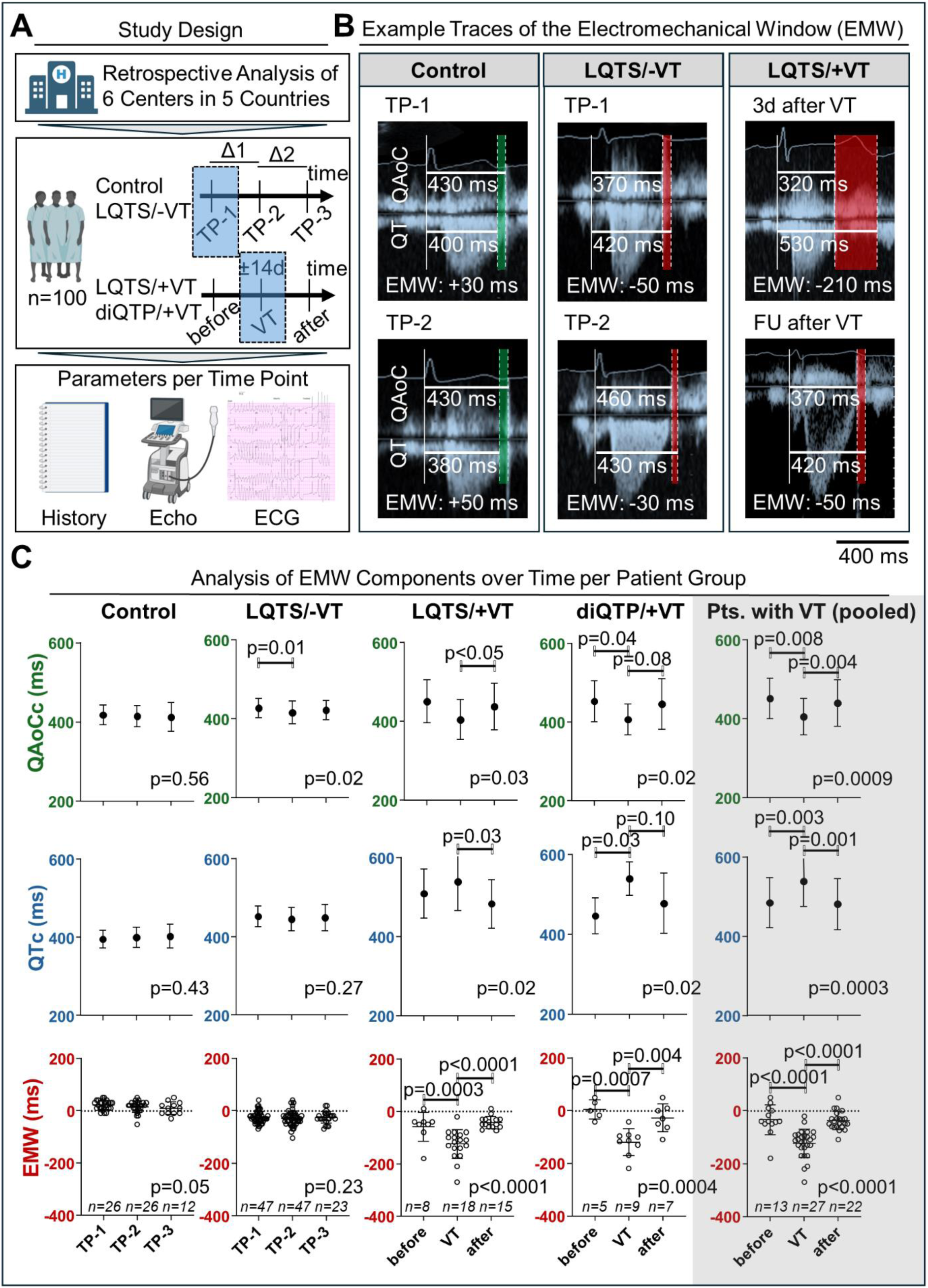
**Temporal Variability of the Electromechanical Window (EMW) and its Components** (A) Study design, follow-up (FU) scheme with up to three study time points (TP), and parameters assessed. Per patient, a minimum of two TPs were assessed, and parameter changes between TPs (ΔQAoCc, ΔQTc, and ΔEMW) were defined as mean of Δ1 and Δ2. The reference time points are highlighted in light blue. (B) For controls, LQTS patients without VT during FU (LQTS/-VT), and LQTS patients with VT during FU (LQTS/+VT), two representative consecutive EMW measurements are depicted. EMW is calculated by subtracting the QT interval (lower line) from the QAoC interval (QRS-onset to aortic-valve closure time; upper line). Note that the EMW is relatively stable in controls and LQTS/-VT, but shows exaggerated negativity in LQTS/+VT patients close to ventricular tachyarrhythmia (VT), and becomes less negative again during FU. (C) Rate-corrected QAoCc and QTc are shown alongside EMW for each patient group. In symptomatic diQTP/LQTS patients, exaggerated EMW negativity close to VT was composed of both a shortened mechanical systole (QAoCc↓) and a prolonged repolarization duration (QTc↑). The pooled analysis discussed in the text is highlighted in light grey. Data are shown as mean±SD.

### QTc, QAoC, and EMW Calculation

The QT time was determined from the single-lead ECG traces (usually lead II), which were recorded during apical aortic-valve Doppler echocardiography, and the tangent method was used to standardize QT assessment.^10^ The QAoC was measured from the onset of the QRS complex to the center of the aortic-valve closure artifact. EMW was calculated by subtracting the QT time from the QAoC of the same heartbeat.^10^ For EMW, no heart-rate correction was performed, as EMW is heart-rate independent.^10,14^ For comparability of QT and QAoC over time, these parameters were rate-corrected using Bazett’s formula. Data were analyzed by one investigator (P.D.), and blindly re-analyzed in part by P.D. and R.t.B., showing high EMW intra-observer (0.99 (0.97-1.0)) and inter-observer (0.99 (0.96-1.0)) agreements.

### Statistical Analysis

Statistical analysis was performed with GraphPad Prism (GraphPad Software Inc., version 10.0.1) and R (version 4.4.1 (2024-06-14)). In Table 1, continuous data are presented as mean ± SD or median (IQR). Statistical comparisons were performed using one-way ANOVA with Tukey’s post-hoc correction or the Kruskal–Wallis test with Dunn’s post-hoc comparison. Binary variables are shown as frequencies and percentages, and an omnibus Fisher’s exact test, followed by pairwise Fisher’s exact test with Bonferroni correction, was used. In Figure 1, a normal distribution of variables was assumed,^15^ and a linear mixed model with random intercept analysis and Tukey post-hoc correction was used. In Figure 2, the association between absolute time-to-event and EMW was assessed based on a linear mixed-effects model with a random intercept. Kruskal-Wallis test with Dunn’s correction was used for comparisons of group Δ’s. Single and multiple logistic regression were based on the ‘glm2’ R-package, and the ‘pROC’ package was used for area under the curve (AUC) and receiver operating characteristic (ROC) analysis. A two-sided p<0.05 was considered statistically significant. Additional details on the methods and statistical analyses are provided in the online supplement.

**Figure 2:**
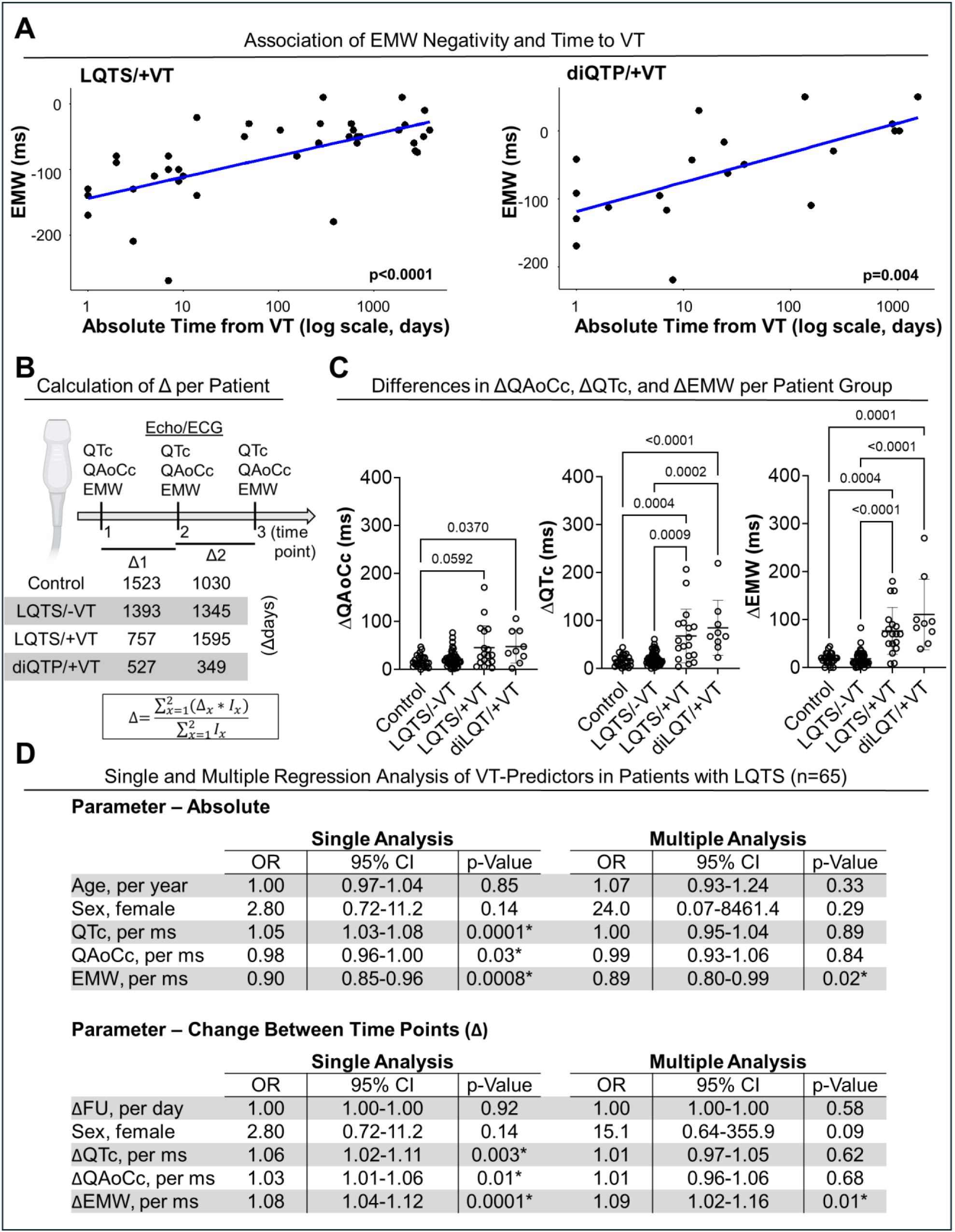
**Time Dependence and Regression Analysis of Electromechanical Parameters** (A) EMW negativity correlates with the absolute time from VT in LQTS and diQTP patients during longitudinal follow-up (FU) in a linear mixed model with a random intercept. (B) In order to determine parameter changes between time points, the ΔQAoCc, ΔQTc, and ΔEMW were calculated according to the depicted formula (‘*I’* being the indicator if the Δ is available for this time interval or not). Mean Δdays are shown for all groups. (C) The analysis of Δ parameters per patient group revealed pronounced dynamicity in patients with VT during FU, especially for ΔQTc and ΔEMW (Kruskal-Wallis test with Dunn’s comparison). (D) Two logistic regression models were created for the analysis of VT predictors in the LQTS cohort, one including the absolute parameter values measured at the reference time points (defined in Table 1), and the other including the mean Δ per parameter measured (defined in Figure 2B). These models only included the data from the LQTS cohort (n=47 LQTS/-VT and n=18 LQTS/+VT patients). The analysis revealed that both the time-point value of QAoCc, QTc, and EMW, as well as the Δ of values of these parameters were associated with the VT status in respective single logistic regression analysis. However, only the EMW and ΔEMW remained significant predictors of VT status in multiple logistic regression analysis.

**Table 1.**
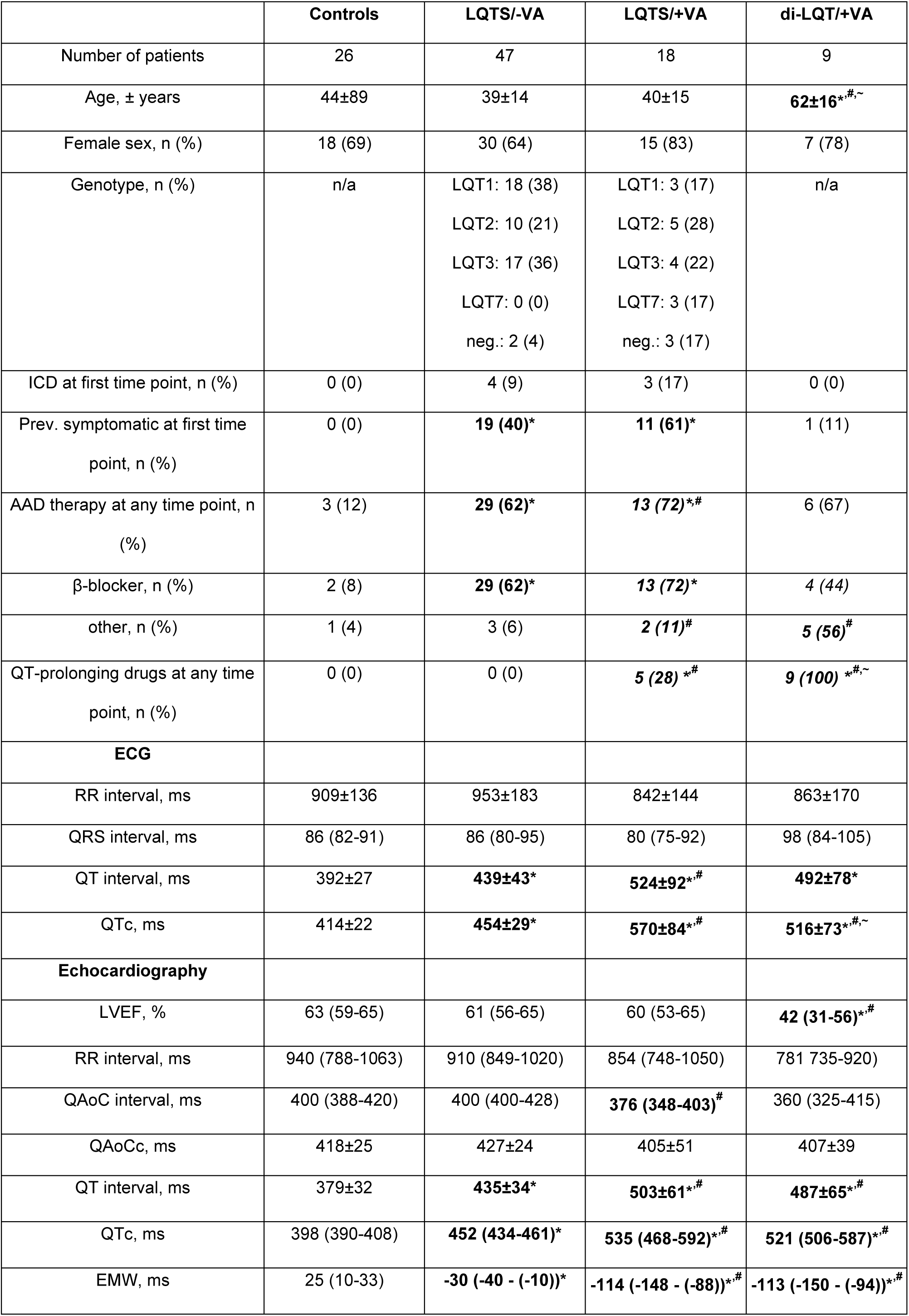
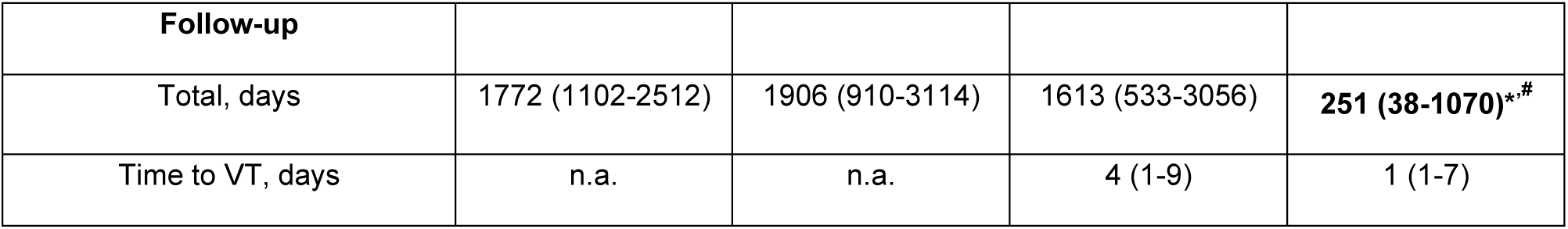
Patient Characteristics. Clinical characteristics of the patients at the reference time point, if not specified otherwise. Data are shown as n (%), mean±SD, or median (25^th^-75^th^ percentile). For controls and LQTS/-VT, the reference is defined as the first echocardiographic evaluation (TP-1 in Figure 1, 2), for LQTS/+VT and diQTP/+VT, the reference is defined as the time point closest to VT (VT in Figure 1,2). For patients with VT during follow-up, time elapsed from the reference point to VT is indicated under follow-up. Continuous data are shown as mean±SD or median (IQR), and one-way ANOVA with Tukey post-hoc test, or Kruskal-Wallis test with Dunn’s comparison were used. Binary variables are shown as frequencies and percentages, and an omnibus Fishers exact test followed by pairwise Fisher’s exact test with Bonferroni correction was used. *: p<0.05 vs. controls, #: p<0.05 vs. LQTS/-VT, ∼: p<0.05 vs. LQTS/+VT. *AAD: antiarrhythmic drug; EMW: electromechanical window; ICD: implantable cardioverter-defibrillator, LQTS: long-QT syndrome, LVEF: left-ventricular ejection fraction, QAoC: QRS-onset to aortic-valve closure time, QAoCc: rate-corrected QAoC by Bazett’s formula, SD: standard deviation, VT: ventricular tachyarrhythmia*.

## Results

### The EMW differs between controls, and LQTS or diQTP patients, depending on symptomatic status

A total of 100 patients and control subjects with a median (IQR) age of 42 (27-57) years and mean±SD follow-up duration of 1869±1360 days were included: 26 controls, 47 LQTS patients without VT (LQTS/-VT), 18 LQTS patients with VT (LQTS/+VT), and 9 diQTP patients with VT (diQTP/+VT). 70% of all patients were female, and LQT1-3 were the most common genotypes in LQTS/-VT (LQT1 38%, LQT2 21%, LQT3 36%, LQT7 0%, genotype-negative 4%) and LQTS/+VT (LQT1 17%, LQT2 28%, LQT3 22%, LQT7 17%, genotype-negative 17%). Overall, beta-blockers were used in 8% of controls (for hypertension), 62% of LQTS/-VT patients, 72% of LQTS/+VT patients, and 44% of diQTP/+VT patients at any time point (Table 1). In the +VT group, 21/27 patients experienced TdP, 3/27 (bidirectional) ventricular tachycardia, and 3/27 VF based on medical records or internal cardioverter-defibrillator (ICD) readouts. EMW at the reference time point was 25 (10 to 33) ms in controls, −33 (−40 to −10) ms in LQTS/- VT, −114 (−148 to −88) ms in LQTS/+VT, and −113 (−150 to −94) ms in diQTP/+VT.

### The EMW is stable in controls and -VT patients, but dynamically rendered more negative in +VT patients close to arrhythmic events

Over time, the EMW remained stable and positive in controls and stable but negative in LQTS/-VT patients (Figure 1B-C). In line with this, QAoCc and QTc remained unchanged between time points. In LQTS/-VT, QAoCc showed a small variation over time with stable QTc, which, however, did not result in significant EMW variability. In contrast, in LQTS/+VT and diQTP/+VT patients, EMW became transiently and significantly more negative when assessed within days before or after VT (Figure 1B- C). In a pooled analysis of all +VT patients, QTc (baseline: 485±63 ms, 555 (1294 to 65) days before VT) had prolonged significantly 2 (1 to 7) days after VT (539±64 ms, p=0.003), and returned to a baseline level during follow-up (482±64 ms, p=0.001), 627 (151 to 2677) days after VT. In the same period, QAoCc (baseline: 452±51 ms) had dropped significantly close to VT (405±46 ms, p=0.008), returning to baseline 440±59 ms (p=0.004) in the follow-up period. Consequently, the EMW (baseline: −35±57 ms) became most negative close to VT (−123±53 ms, p<0.0001), returning to baseline values at follow-up (−38±35 ms, p<0.0001). When analyzing the LQTS/+VT and diQTP/+VT study groups separately, both showed similar changes in the electrical and mechanical components of EMW near the time of VT (see Online Supplement for details). Also, at time points remote from VT, LQTS/+VT patients had more negative EMW values than LQTS/-VT patients (before VT (−584 (−1543 to −212) days): −45 (−73 to −40) vs. −30 (−40 to −10) ms, p=0.009, follow-up after VT (+722 (276 to 2864) days): −50 (−60 to −30) vs. −30 (−40 to −10) ms, p=0.003). Temporal EMW variability was similar for LQTS patients with LQT1-3, Andersen-Tawil syndrome (LQT7), or genotype-negative patients. Interestingly, absolute time from VT and EMW negativity were associated significantly for LQTS/+VT (p<0.0001) and diQTP/+VT patients (p=0.004) (Figure 2A).

### EMW restitution after VT is independent of beta-blocker therapy

In LQTS/+VT patients (n=4) not treated with beta-blockers before the event *nor* during follow-up, QAoCc was significantly shorter close to VT than at follow-up (391±34 vs. 416±34 ms, p=0.03). QTc was numerically longer close to VT than during follow-up (505±54 vs. 463±49 ms, p=0.12), and consequently, the EMW was more negative close to VT than during follow-up (−110±26 vs. −43±14 ms, p=0.01). Thus, EMW normalization during follow-up also occurred in the absence of beta-blocker therapy.

In the next step, we compared the four beta-blocker-naive LQTS/+VT patients to seven LQTS/+VT patients who were initiated on beta-blocker therapy after the arrhythmic event (5/7 propranolol, 2/7 metoprolol). QAoCc, QTc, and EMW were similar between groups close to VT and during follow-up (two-way repeated measures ANOVA with Sidak correction, see Online Supplement and Supplementary Figure 2).

### Absolute EMW and EMW dynamics (ΔEMW) are independently associated with VT status in LQTS

Mean QAoCc, QTc, and EMW changes between time points (Δ) were quantified for every patient according to the formula shown in Figure 2B. In general, ΔQTc and ΔEMW were significantly larger in LQTS/+VT and diQTP/+VT patients than in controls or LQTS/-VT patients (Figure 2B,C). +VT patients of both LQTS and diQTP groups showed similar ΔQAoCc (29 (14 to 81) vs. 38 (21 to 80) ms, p=0.99), ΔQTc (53 (25 to 94) vs. 67 (50 to 106) ms, p=0.99), and ΔEMW (70 (48 to 93) vs. 92 (63 to 145) ms, p=0.99, respectively, Figure 2C). A single logistic regression model (Figure 2D) for the LQTS/-VT and LQTS/+VT patients (n=65) revealed that EMW (p=0.0008), QTc (p=0.0001), and QAoCc (p=0.03) were associated with VT status, but only EMW (p=0.02) remained significant after multiple regression analysis. In a second model investigating parameter changes (Δ) over time, ΔEMW (p=0.0001), ΔQTc (p=0.003), and ΔQAoCc (p=0.01) were predictors of VT status, but only ΔEMW (p=0.01) remained significant after multiple regression analysis (Figure 2D). In a ROC analysis for identifying LQTS/+VT patients, an area under the curve (AUC) of 0.97 (confidence interval (CI): 0.90-1.00) for EMW was found, with a cut-off EMW of −75 ms (sensitivity: 0.94, specificity: 1.00). For ΔEMW, an AUC of 0.88 (CI: 0.74-0.99) was found, with a cut-off of −39 ms (sensitivity: 0.83, specificity: 0.96).

### Representative cases with a high arrhythmic burden illustrate temporarily accentuated EMW negativity preceding or shortly following VT

In an LQT3 patient with VT storm and a history of major adverse cardiovascular events, TdP onset was coincidentally recorded with ECG-echocardiography, revealing a markedly negative EMW of −160 ms in the last sinus-rhythm beat before TdP (Figure 3A).^10^ Years later, this patient experienced two appropriate ICD shocks for TdP, each coinciding with a transient exaggeration of EMW negativity. During clinically-stable periods, EMW was consistently less negative at −50 ms (Figure 3A).

**Figure 3:**
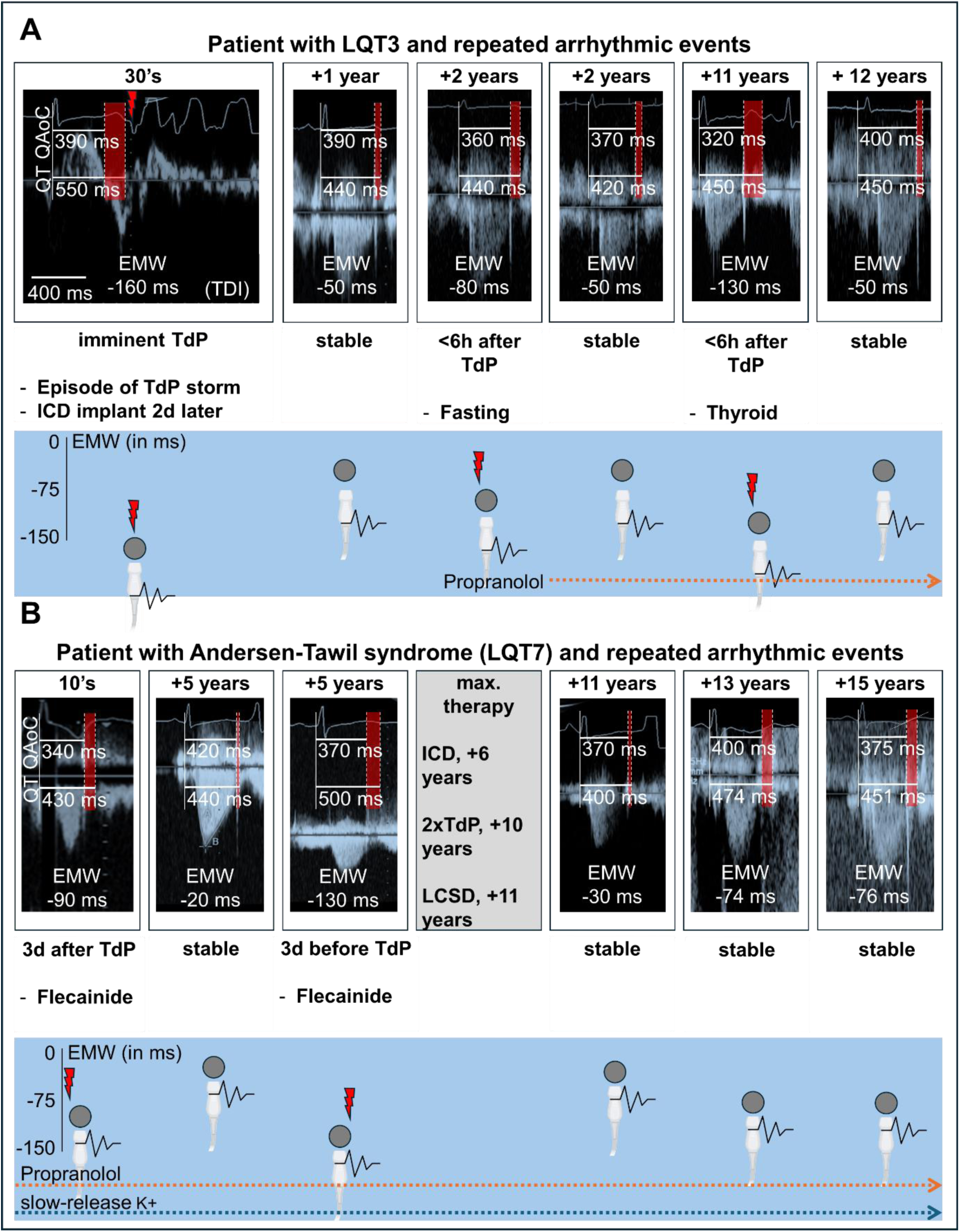
**Examples of EMW Temporal Variability Around Moments of Torsades de Pointes in Highly Symptomatic LQTS Patients** (A) In this LQT3 patient in their thirties, TdP storm occurred, and TdP onset was coincidentally recorded with tissue-Doppler echocardiography (TDE), revealing a markedly-negative EMW right before TdP induction. During follow-up of >12 years, the patient experienced two adequate ICD shocks in the context of a fasting period and hyperthyroidism, both coinciding with transient exaggeration of EMW negativity measured on the day of the event. (B) In this teenager LQT7 patient, flecainide therapy was initiated twice for premature ventricular complex (PVC)-suppression. Flecainide therapy led to a transient exaggeration of EMW negativity, and polymorphic VT/TdP occurred. Interestingly, a markedly negative EMW was found three days before TdP occurred. Grey circles indicate EMW value at time of measurement; ECG/echo icons indicate points of EMW assessment, while the lightning icon marks the occurrence of TdP/VT relative to ECG-echocardiography. Both examples demonstrate that accentuated EMW negativity develops prior to VT and is not a post-hoc phenomenon occurring in the aftermath of severe arrhythmia.

In a case of a symptomatic LQT7 patient with a high burden of premature ventricular complexes (PVCs) and repeated episodes of non-sustained VT, flecainide therapy was started. The patient was admitted shortly thereafter with recurrent episodes of TdP, prompting flecainide discontinuation. The EMW was assessed three days later (−90 ms), and returned to more positive values during follow-up (Figure 3B). Several years later, ajmaline provocation testing demonstrated effective PVC suppression, and flecainide was restarted. One month later, routine ECG-echocardiography showed an LVEF of 45%, with again a markedly negative EMW (−130 ms). Three days afterwards, the patient developed polymorphic VT and TdP, leading to the discontinuation of flecainide and ICD implantation. After two additional arrhythmic events years later, the patient underwent left cardiac sympathetic denervation (LCSD), which led to a substantial reduction in PVC burden and complete suppression of TdP. Since then, EMW values remained stably negative, and did not drop further. The patient has remained free from TdP or any sustained VT, and LV ejection fraction remained >50%, thereafter (Figure 3B).

## Discussion

In the setting of ventricular repolarization prolongation, TdP occurs under dynamic rather than static conditions in which electrical instability and emergent ventricular ectopy precede sustained arrhythmia.^16,17^ This study demonstrates that transient mechanical changes (QAoCc↓) parallel electrical alterations (QTc↑), and mark pending VT in LQTS and diQTP patients. Consequential temporary accentuation of EMW negativity is an independent marker for imminent VT in LQTS patients, and is superior to QTc or its long-term variability (ΔQTc). These findings suggest that periodic non-invasive EMW monitoring to detect these changes provides significant added value for arrhythmia-risk stratification beyond the repeated determination of QTc.

Mechanical abnormalities in LQTS syndrome patients were first described by Nador et al. in the 1990s, who observed an abnormal systolic contraction phase in LQTS patients,^18^ which was attributed to abnormal intracellular Ca^2+^ handling.^19,20^ Also, an abnormal relation between electrical and mechanical systole was found in LQTS patients, which was accentuated by exercise testing, indicating the central role of the autonomic nervous system.^21^ Apart from the overall EMW assessment,^11,12^ exercise-induced variability in EMW has been observed in LQTS patients, particularly in those who were previously symptomatic.^22^

The interplay of multiple (partially concealed) factors that align at a critical moment to provoke arrhythmic events may also lead to accentuated EMW negativity. While acute drivers of arrhythmic instability are often unknown in congenital LQTS patients, drug-mediated I_Kr_ block serves as a dominant factor in diQTP patients, whether or not against a background of repolarization lability by other causes.^23^ To improve arrhythmia prediction in patients with LQTS and diQTP, the change of EMW (ΔEMW) around moments of TdP was investigated. Our analysis demonstrates that ΔEMW is an independent predictor of imminent arrhythmia risk, with an optimal cut-off of −39 ms over time, yielding a sensitivity of 0.83 and a specificity of 0.96. Interestingly, LQTS and diQTP patients with pending TdP, VF and/or bidirectional VT showed remarkable similarity in the temporal variability of the EMW and its components. This suggests that the mechanisms driving EMW negativity and arrhythmogenesis partly overlap. These drivers likely involve electromechanical reciprocity, meaning that transient changes in both electrics and mechanics can influence each other.^2^ For example, early and delayed afterdepolarizations may lead to mechanical abnormalities such as post-systolic shortening, aftercontractions, and dispersion of contraction,^24,25^ whereas primary mechanical abnormalities (e.g., acute afterload increases during exercise) may, in turn, precipitate repolarization instability in susceptible myocardium.^2,26^ Additionally, episodes of increased sympathetic activity and pause-dependent potentiation may transiently accentuate electromechanical dispersion. Such episodes may vary between individuals, as exemplified by inter-individual differences in adrenergic responsiveness and neural regulation of the heart, even among patients harboring the same pathogenic gene mutation.^27^ Experimental work in a canine drug-induced LQT1 model showed that left-stellate ganglion stimulation led to accentuated EMW negativity (and TdP initiation) due to divergent autonomic effects on electrics (QT) and mechanics (QAoC).^10,28^ Conversely, beta-blocker treatment and left-stellate ganglion denervation rendered EMW less negative in LQTS patients, reducing TdP occurrence.^10,11,29^ Our results suggest that EMW normalization during follow-up cannot be solely attributed to the initiation of beta-blocker therapy after ventricular arrhythmia, as a comparable reduction in EMW negativity was observed in beta-blocker-naïve patients, albeit with different absolute values. Furthermore, electrolyte imbalance (e.g., hypokalemia, hypomagnesemia) can impair electromechanical stability.^1,2^ Finally, abnormal gene regulation, or proarrhythmic drug effects influencing ion-channel function and/or trafficking, may further aggravate electromechanical instability in susceptible patients, ultimately facilitating TdP. ^23,30^

In the present study, EMW was assessed within 2 weeks of the arrhythmic event (median 2 (1 to 7) days to arrhythmia), as ECG-echocardiography was typically performed during the clinical work-up following TdP. This raises two questions: 1) Is EMW negativity a cause or consequence of ventricular arrhythmia? 2) What is the time scale of the EMW change? To address the first issue, we have carefully reviewed cases with ECG-echocardiography just before the occurrence of VT (Figure 3), revealing that pronounced EMW negativity *precedes* the arrhythmia, confirming experimental data.^12,26^ Therefore, we argue that accentuated EMW negativity adds to the substrate for TdP. Regarding the second question, changes in autonomic tone may modulate the EMW within seconds to minutes,^22,31^ whereas myocardial remodeling associated with disease progression influences EMW over longer time scales.^32,33^ Although unknown for LQTS and diQTP, significant EMW negativity was already observed 79 (21-192) days before VT occurred in ICD carriers with mostly ischemic or dilated cardiomyopathy.^33^

Ultimately, the findings of the present study raise the question of whether – and, if so, which – preventive measures should be implemented when a large EMW drop is found in a LQTS patient or other susceptible patients. While prospective trials are needed to answer this question, it seems reasonable to speculate that this finding should trigger a thorough investigation of potential underlying and addressable causes (e.g., electrolyte imbalance, new (QT-prolonging) medication, insufficient beta-blocker adherence). Additionally, prospective trials should investigate which methods of continuous EMW monitoring (e.g., photoplethysmography combined with repolarization monitoring) provide further insights into electromechanical dynamics under various conditions.^10^

## Limitations

The patient number in this retrospective study was relatively small and follow-up intervals varied between subjects, limiting the assessment of the speed of change of the parameters, which would be available in a prospective study. However, as repeated ECG-echocardiographic analyses are not routinely performed in LQTS patients in most clinics, our dataset provides unique insights into the temporal variability of EMW.

## Conclusions

While temporal stability characterizes the EMW of control subjects and asymptomatic LQTS patients, EMW negativity is temporarily accentuated in LQTS/diQTP patients with pending VT. Non-invasive EMW monitoring outperforms QTc in detecting recent ventricular arrhythmia episodes in patients with LQTS and diQTP, potentially improving imminent arrhythmia risk prediction and patient management.

## Funding

PD was supported by the German Academic Scholarship Foundation and the German Research Foundation (Walter Benjamin Programme, 529532291). AKR was supported by Heidelberg University Medical Faculty’s Olympia Morata Grant. MEM was funded by the Career Development Programm – Short Term Fellowship – of Heidelberg University, Faculty of Medicine. MS is supported by the Dutch Cardiovascular Alliance, an initiative with support of the Dutch Heart Foundation, and Stichting Hartedroom for financing the Double Dose program 2020-B005. SH receives personal fees for independent scientific advice on early development in the field of heart failure for AstraZeneca, Ribocure, and CSL Behring, and receives research support from AstraZeneca and CSL Behring; the research leading to these results has received funding from the European Union Commission’s Seventh Framework program under grant agreement N° 305507 (HOMAGE). The support of IMI2-CARDIATEAM, from the Innovative Medicines Initiative 2 Joint Undertaking (JU) under grant agreement N° 821508 is acknowledged; The JU receives support from the European Union’s Horizon 2020 research and innovation program and EFPIA; the support from the Netherlands Cardiovascular Research Initiative, an initiative with support of the Dutch Heart Foundation, Dutch Cardiovascular Alliance Double Dosis, 2020-B005; ZonMW-Metacor. MR was supported by a grant from the “Gottfried and Julia Bangerter-Rhyner-Stiftung” Switzerland and a grant from the University Hospital of Bern, Inselspital (“Nachwuchsförderungs-Grant”). KEO received funding by a grant from the Swiss National Science Foundation (SNF 310030_197595) and a Bern Center of Precision Medicine Lighthouse grant. PV received grants from The Netherlands CardioVascular Research Initiative (CVON 2017-13 VIGILANCE and CVON 2018-30 PREDICT2), Den Haag, The Netherlands, and the Health Foundation Limburg, Maastricht. RtB received grants from The Netherlands Organization for Scientific Research (Veni grant, 0915016181013), and the Health Foundation Limburg, Maastricht.

## Data Availability

Data will be shared by the corresponding author upon reasonable request and after legal assessment.

## Acknowledgements

The authors wish to express their gratitude to Prof. Dr. Christian Sohns MD (Herz- und Diabeteszentrum Nordrhein-Westfalen, Bad Oeynhausen, Germany) and Simone van Wanroij-Jorissen RN (Maastricht University Medical Center +) for their valuable contributions to the study.

## Disclosure of Interest

None

## Data availability statement

Data will be shared upon reasonable request and after legal assessment by the corresponding author (R.t.B.).

## Abbreviations

AUC: Area Under the Curve
diQTP: Drug-Induced Long-QT Syndrome
EMW: Electromechanical Window
ICD: Internal Cardioverter-Defibrillator
LCSD: Left Cardiac Sympathetic Denervation
LQTS: Long-QT Syndrome
QAoC: Aortic-Valve Closure Time
ROC: Receiver Operating Characteristic
SCD: Sudden Cardiac Death
TdP: Torsades de Pointes
VF: Ventricular Fibrillation
VT: Ventricular Tachyarrhythmia

## Online Supplement

### Methods

#### Study Design and Criteria

Patients were prospectively and retrospectively included between 09.2023 and 10.2024 from six centers in five European countries (Maastricht (NL), Tel Aviv (ISR), Bern (CH), Heidelberg (DE), Innsbruck (AU), and Bad Oeynhausen (DE)). All patients were ≥18 y.o. at the time of study inclusion, and provided written informed consent according to institutional regulations.

Control patients were recruited after serious cardiac conditions were excluded. This was defined as the absence of: dialysis, cardiomyopathy (LVEF <50%), LV hypertrophy (IVSd >11 mm), diastolic heart failure, moderate to severe valvular disease, mitral valve prolapse, coronary artery disease, AV block, sick sinus syndrome, and the use of QT prolonging medication (according to https://crediblemeds.org/).

LQTS was defined according to the ESC guidelines^1^, and diLQT was defined as patients with transient QT prolongation during treatment with known repolarization-delaying drugs (https://crediblemeds.org/). Patient characteristics were assessed at two or three time points, and arrhythmic events (=VT) during follow-up were defined as documented and symptomatic (non-sustained) torsade de pointes, ventricular tachycardia, or ventricular fibrillation. In case of VT, patients were only included if an echo and ECG were performed within 14 days of VT. Patients from either group were only included if a minimum of two timepoints with complete data were available.

Sample size calculation (G*Power 3.1, Düsseldorf, DE) was based on a pilot dataset (n=30 patients; 10 controls, 10 LQTS/-VT, 5 LQTS/+VT, 5 diLQT/+VT), and used the EMW change between time points (ΔEMW) per patient as the input variable. An effect size *f* = 0.71 was determined, and for the fixed-effects omnibus one-way ANOVA a significance level (α) of 0.05 and a power of 0.80 were specified. This calculation indicated that a total of 28 patients (7 patients per group) was needed for an adequately powered analysis.

Intra- and interobserver variability assessment of the EMW measurements was performed with RStudio (version 4.4.1 (2024-06-14)), and the ‘irr’ package was used with a two-way model and an agreement type as settings. Intraobserver agreement was high, with 0.92 (0.79-0.97) for QAoC, 0.96 (0.91-0.99) for QTc, and 0.99 (0.97-1.0) for EMW. Interobserver agreement was also high, with 0.92 (0.79-0.97) for QAoC, 0.96 (0.90-0.99) for QT, and 0.99 (0.96-1.0) for EMW.

#### Statistical Analysis

Statistical analysis was performed with GraphPad Prism (GraphPad Software Inc., version 10.0.1) and R (version 4.4.1 (2024-06-14)).

In Table 1, continuous data are shown as mean±SD or median (IQR), and one-way ANOVA with Tukey post-hoc correction, or Kruskal-Wallis test with Dunn’s comparison were used. Data distribution was assessed with the Kolmogorow-Smirnow test. Binary variables are shown as frequencies and percentages, and an omnibus Fishers exact test followed by pairwise Fishers exact test with Bonferroni correction was used.

In Figure 1, normal distribution of variables was assumed as linear-mixed models are robust towards violations of distributional assumptions^2^. This approach was chosen, as the sample sizes in some groups were rather small and the dataset may therefore not be normally distributed due to sample characteristics and not due to true non-normal distribution. Data are shown as mean±SD, and a linear mixed model with a random intercept analysis (‘tidyverse’, ‘lmerTest’, and ‘emmeans’ R-packages) with Tukey post-hoc correction was used. The degrees of freedom were assessed by the Satterthwaite method.

In Figure 2, association between absolute time-to-event and EMW was assessed with a linear mixed-effects model with a random intercept (‘nlme’ and ‘tidyverse’ R-package). Due to non-normal distribution, Kruskal-Wallis test with Dunn’s correction was used for between group comparisons of group Δ’s in Figure 2B. Single and multiple logistic regression was based on the ‘glm2’ R-package, and the ‘pROC’ package was used for area under the curve (AUC) and receiver operating characteristic (ROC) analysis. AUC was calculated using the bootstrap method with a boot.n of 1000. The EMW and ΔEMW cut-off for the values of highest sensitivity and specificity were determined using Youden’s J statistic (max(sensitivities+specificities)) within the ‘pROC’ package.

A two-sided p<0.05 was considered to be statistically significant.

R package versions used:

‘tidyverse’ – version 2.0.0

‘lmerTest’ – version 3.1.3

‘emmeans’ – version 1.10.3

‘nlme’ – version 3.1.164

‘glm2’ – version 1.2.1

‘pROC‘ – version 1.18.5

‘ggplot2‘ – version 3.5.1

### Results

#### EMW Differences in Patients without VT during Follow-up

Within the group of 47 LQTS/-VT patients, n=28 were completely asymptomatic prior to inclusion in this study, and n=19 previously experienced symptoms (e.g., syncope, palpitations, or (NS)VT).

Interestingly, although the QTc (448±24 vs. 456±29 ms, p=0.33) and QAoCc (430±21 vs. 422±29 ms, p=0.27) did not significantly differ between previously asymptomatic and previously symptomatic patients in the LQTS/-VT group, the EMW was less negative in asymptomatic LQTS/-VT patients than in previously symptomatic LQTS/-VT patients (−17±21 vs. −33±19 ms, p=0.01), thus indicating that also symptom status beyond sustained VT during follow-up corresponds to a more negative EMW.^3,4^ See Supplementary Figure 1.

Temporal Variability of the EMW and its Components in LQTS/+VT and diLQT/+VT:

In LQTS/+VT patients, the baseline QAoCc −584 (−1543 to −212) days before the arrhythmic event was 451±55 ms, and was numerically rendered shorter to 405±51 ms (p=0.13) when measured 4 (1 to 9) days after VT. During the follow up measurement 722 (276 to 2864) days after VT, the QAoCc became longer again (438±59 ms, p=0.047). Opposite to that, the QTc (baseline 510±62 ms) was rendered numerically longer to 539±73 ms close to VT (p=0.11), but returned to a lower level during follow up (483±61 ms, p=0.03). As a result, the baseline EMW (−59±55 ms) became significantly more negative close to ventricular arrhythmia (−125±56 ms, p=0.0003). A normalization of the EMW towards more positive values (−43±23 ms, p<0.0001) was found in the follow up measurements.

In diLQT/+VT patients, the baseline QAoCc −24 (−1294 to −13) days before the arrhythmic event was 453±52 ms, and was significantly rendered shorter to 407±39 ms (p=0.04) when measured 1 (1 to 7) days after VT. During the follow up measurement 156 (37 to 894) days after VT, the QAoCc tended to become longer again (446±64 ms, p=0.08). Opposite to that, the QTc (baseline 446±45 ms) was rendered longer to 539±42 ms close to VT (p=0.03), but tended to return to a lower level during follow-up (478±76 ms, p=0.10). As a result, the baseline EMW (4±37 ms) became significantly more negative close to ventricular arrhythmia (−120±51 ms, p=0.0007). A normalization of the EMW towards more positive values (−28±53 ms, p=0.004) was found in the follow-up measurements.

We also compared LQTS/+VT patients that were not treated with beta-blockers close to VT and during follow-up (n=4, LQTS/+VT/-BB) with LQTS/+VT patients that had no beta-blocker therapy close to the event, but beta-blocker therapy during follow-up (n=7, LQTS/+VT/+BB). There was no difference in QTc (505±54 vs. 535±65 ms, p=0.67), QAoCc (391±34 vs. 382±35 ms, p=0.94), and EMW (−110±26 vs. −136±50 ms, p=0.44) close to the arrhythmic event between -BB and +BB LQTS/+VT patients, both beta-blocker naïve at this point. Also during follow-up (after beta-blocker therapy was started in the +BB group), QTc (463±49 vs. 476±58, p=0.92), QAoCc (416±34 vs. 437±61, p=0.71), and EMW (−43±14 vs. −36±29 ms, p=0.94) did not differ between -BB and +BB LQTS/+VT patients, thus indicating that EMW normalization during follow-up cannot only be subjected to beta-blocker therapy initiation. See Supplementary Figure 2.

**Supplementary Figure 1:**
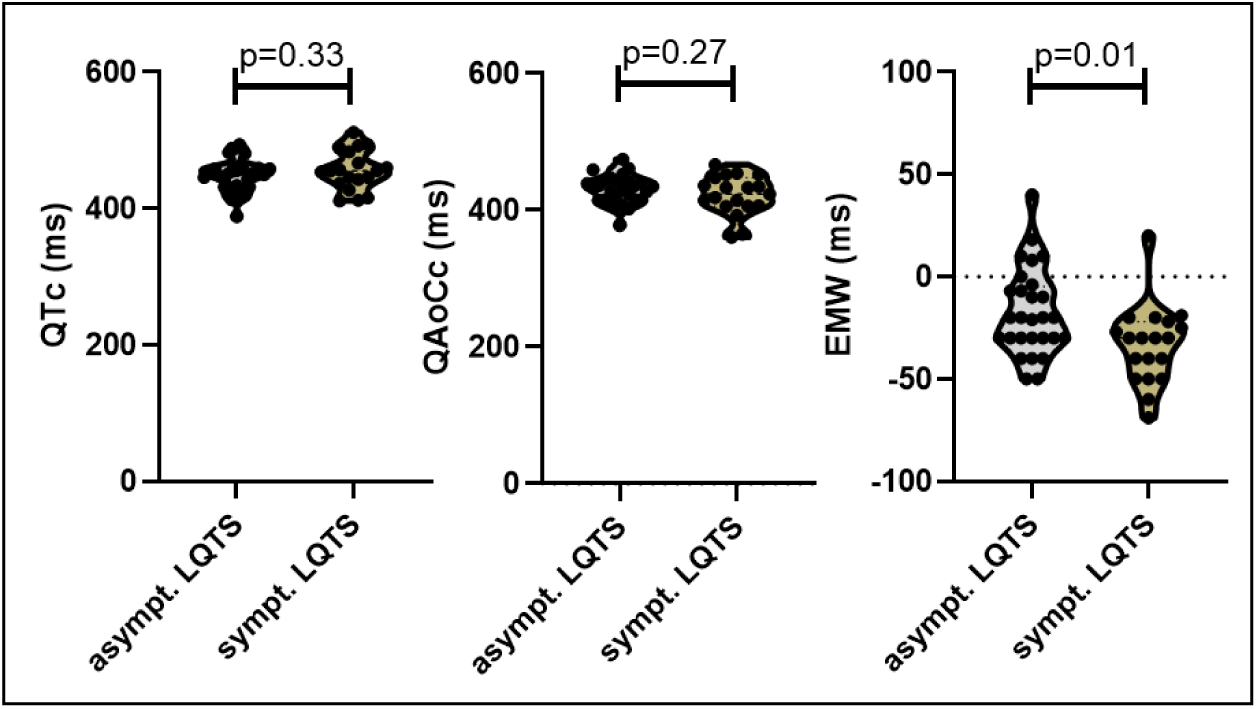
EMW differences between asymptomatic (asympt.) and symptomatic (sympt.) LQTS patients without VT (LQTS/-VA) during follow-up.

**Supplementary Figure 2:**
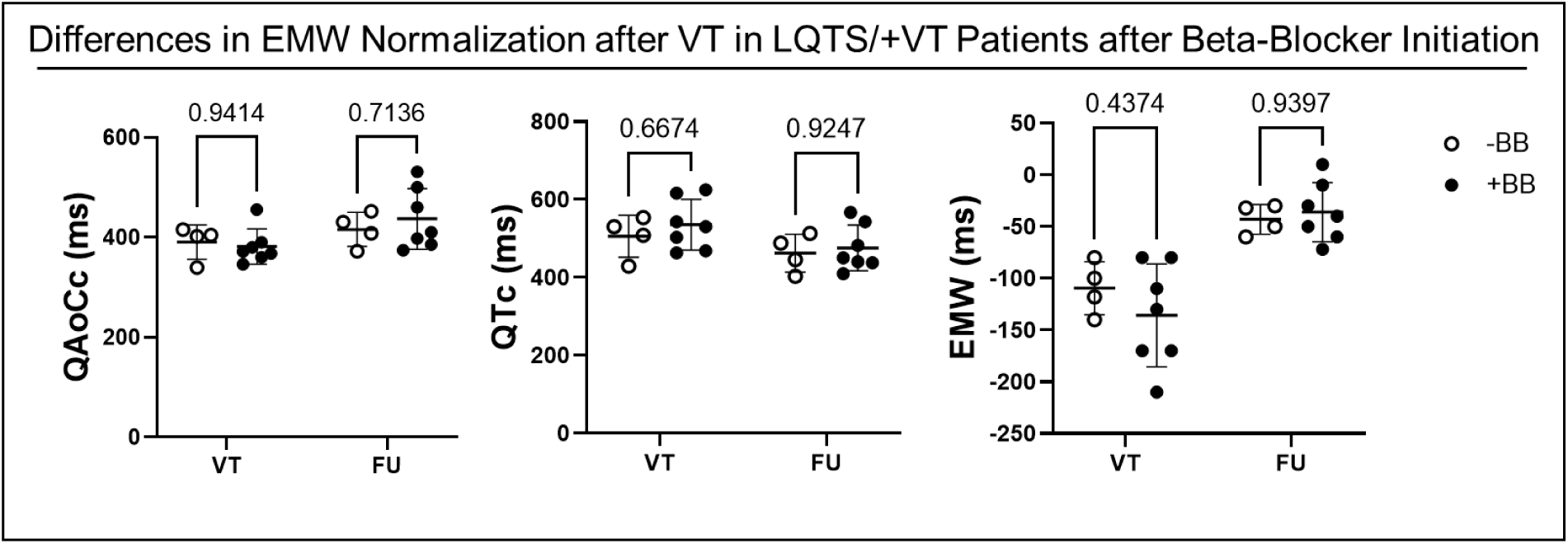
EMW dynamics after VT and its relation to the initiation of beat-blocker therapy in LQTS/+VT patients. EMW and its components adapted similarly during follow-up after VT, both in patients without initiation of beta-blockers (-BB) and in patients with initiation of beta-blocker therapy (+BB). All patients were beat-blocker naïve during the arrhythmic event.

## Notes

### Competing Interest Statement

The authors have declared no competing interest.

### Author Declarations

1. Maastricht University Medical Center: Medisch-ethische Toetsingscommissie Maastricht UMC+ T.a.v. METC azM/UM P. Debyelaan 25 Postbus 5800 6202 AZ Maastricht The Netherlands status/decision: this commission gave ethical approval for this work 2. Heidelberg University Medical Center: Ethikkommission I Heidelberg Alte Glockengiesserei 11/1 69115 Heidelberg Germany status/decision: this commission gave ethical approval for this work 3. Sheba Medical Center: IRB-Helsinki Committee 2 Derech Sheba Tel Hashomer, Ramat Gan 52621/52662 Israel status/decision: this commission gave ethical approval for this work 4. Herz- und Diabeteszentrum NRW Ethikkommission der Medizinischen Fakultaet der Ruhr-Universitaet Bochum Sitz Ostwestfalen Georgstr. 11 32545 Bad Oeynhausen Germany status/decision: this commission gave ethical approval for this work 5. Bern University Medical Center / Inselspital: Gesundheits-, Sozial-und Integrationsdirektion des Kantons Bern (GSI) Kantonale Ethikkommission Murtenstrasse 31 Hoersaaltrakt Pathologie, Eingang 43A, Buero H372 3010 Bern Switzerland status/decision: this commission gave ethical approval for this work 6. Innsbruck University Medical Center: Geschaeftsstelle der Ethikkommission der Medizinischen Universitaet Innsbruck Haus 11 Vinzenzgebaeude 4 OG Eingang A Postanschrift: Anichstrasse 35 A-6020 Innsbruck Austria status/decision: this commission gave ethical approval for this work

